# Moderate Kidney Dysfunction Independently Increases Sudden Cardiac Arrest Risk: A Community-Based Study

**DOI:** 10.1101/2025.03.12.25323871

**Authors:** Thien Tan Tri Tai Truyen, Audrey Uy-Evanado, Harpriya Chugh, Kyndaron Reinier, David M. Charytan, Angelo Salvucci, Jonathan Jui, Sumeet S. Chugh

## Abstract

**Background:** Moderate kidney dysfunction is independently associated with increased cardiovascular mortality. Sudden cardiac arrest (SCA) accounts for at least 25% of chronic kidney disease (CKD) mortality.

**Methods:** We conducted a case-control study within an ongoing, prospective, community-based investigation of out-of-hospital SCA in the Portland, Oregon, metropolitan area (population ∼ 1 million) from February 1st, 2002, to December 31st, 2020. Analysis included individuals aged 40 to 75 who experienced SCA (cases) and individuals with no history of SCA (controls), with creatinine levels measured prior to SCA/enrollment. Moderate CKD was defined by an estimated glomerular filtration rate (eGFR) of 30 to <60 mL/min/1.73 m^2^ (2021 CKD-EPI formula). A population-based SCA study in Southern California was used for validation.

**Results:** We compared 2,068 SCA cases and 852 controls (mean ages: 61.4±8.5 and 62.7±8.0 years; males: 69.9% and 67.4%). SCA cases had more moderate CKD (17.7% vs. 14.7%, p<0.001) and lower eGFR (74.7 vs. 80.9 mL/min/1.73 m^2^, p<0.001) than controls. Multivariable regression demonstrated that moderate CKD was an independent risk factor for SCA (OR: 1.33, 95% CI: 1.03-1.72). Each 10 mL/min/1.73 m^2^ eGFR drop below 90 increased SCA risk (OR: 1.24, 95% CI: 1.18–1.31). Similar findings were observed in the validation cohort (817 SCA and 3,249 controls), where moderate CKD was associated with SCA (OR: 1.51, 95% CI: 1.16–1.97).

**Conclusion:** Moderate CKD is associated with an increased risk of SCA in the general population. Further research into the potential integration of moderate renal dysfunction into SCA risk stratification are warranted.

**Clinical Perspective:**

- Our findings indicate that even moderate renal dysfunction was independently associated with sudden cardiac arrest (SCA) in two geographically distinct populations. A decline in eGFR below 90 mL/min/1.73m^2^ exhibited a dose-response relationship with SCA.
- Among SCA cases, moderate CKD was linked to a higher likelihood of presenting with a non-shockable rhythm at the time of the even, and lower survival rates to hospital discharge compared to those with normal or mild CKD.
- The findings have implications for the potential integration of moderate renal dysfunction and specifically eGFR, given its dose-response relationship, into SCA clinical risk stratification.

## Introduction

Sudden cardiac death (SCD) accounts for a significant proportion of mortality (up to 25%) among individuals with chronic kidney disease (CKD), particularly in those with advanced CKD.^1–3^ Associations between moderate CKD, defined as an estimated glomerular filtration rate (eGFR) of 30–60 mL/min/1.73 m^2^,^4^ and SCD have been described but remain controversial.

Previous studies have reported that moderate CKD is associated with an increased risk of SCD in individuals with left ventricular dysfunction or coronary artery disease.^5,6^ A study in the general population reported that non-dialysis-dependent CKD patients face an elevated risk of SCD and worse outcomes after cardiac arrest, with survival rates decreasing as GFR declines.^7^ However, other evidence suggests no significant association between estimated GFR (eGFR) levels of 40– 60 mL/min/1.73 m^2^ and SCD compared to eGFR levels above 60 mL/min/1.73 m^2^).^8^ Moreover, due to the small number of SCD cases in previous studies, individuals with moderate CKD have often been grouped with those with advanced CKD for analysis, resulting in a lack of sufficient investigation into moderate CKD as a distinct category.^6–8^ Therefore, there is a need for studies specifically designed to evaluate the relationship between moderate CKD, as a distinct condition, and SCD, as well as to investigate the potential underlying mechanisms driving this association in this unique patient population.

Additionally, in previous studies, eGFR was commonly calculated using either the Modification of Diet in Renal Disease (MDRD-4) formula or the Chronic Kidney Disease Epidemiology Collaboration (CKD-EPI) 2009/2012 formula, both of which incorporate patient age, sex, serum creatinine levels, and race. However, these equations have been shown to yield higher eGFR values for individuals identified as Black compared to non-Black individuals with similar characteristics.^9^ Lesley et al. proposed a new equation in 2021 for estimating GFR that excludes race as a variable.^10^ This approach was subsequently endorsed by the National Kidney Foundation –American Society of Nephrology Task Force. Recent studies indicate that applying the updated CKD-EPI 2021 formula has led to a significant reclassification of CKD patients, impacting how disease severity and progression are assessed.^11,12^ Moreover, moderate CKD, classified based on eGFR calculated from serum creatinine using the CKD-EPI 2012 or MDRD-4 formula, has been reported to predict SCD in some studies, while other studies have found no such association, particularly in older patients or the general population.^6,7,13–15^

We conducted this study to address these knowledge gaps and to evaluate how moderate CKD, assessed using the updated CKD-EPI 2021 criteria, influences the risk of sudden cardiac arrest (SCA) at the community level. Additionally, our study aims to investigate potential disparities in resuscitation outcomes across the CKD severity spectrum in SCA cases and highlight the potential mechanisms linking moderate CKD and SCA.

## Methods

### Data availability statement

The data in this manuscript are from an ongoing study, and there is currently no IRB-approved mechanism by which this data will be deposited in a public repository. All analytical methods are included in this published article. De-identified participant data will be made available after publication upon reasonable request to the corresponding author, following approval of a proposal and a signed data use agreement.

### Study population

Study participants were selected from the Oregon Sudden Unexpected Death Study (ORSUDS), an ongoing prospective, population-based study that enrolls individuals experiencing out-of-hospital SCA within the Portland, Oregon metropolitan area (catchment population approximately 1 million). Cases were identified through multiple sources, including first responders (Portland Fire Department and local ambulance services), the county medical examiner’s office, and emergency departments of participating hospitals. The study methods have been detailed in prior publications.^16^ Briefly, after receiving the SCA subject’s information, a comprehensive review of each case was conducted, encompassing circumstances of the event, clinical history, and, when available, autopsy findings. Cases of SCA with a likely cardiac etiology were determined through an adjudication process involving three physicians. SCA was defined as a sudden, pulseless collapse due to a likely cardiac etiology, occurring rapidly after symptom onset when witnessed or if unwitnessed, within 24 h of the subject being last seen in their usual state of health.^17^ Cases with non-cardiac etiologies, such as trauma, drowning, overdose/substance abuse, terminal illness, malignancies not in remission, or extracardiac causes (e.g., pulmonary embolism), were excluded.

Controls for ORSUDS were prospectively recruited from the same geographic region and time frame as the cases, ensuring a population with a medium-high risk profile, marked by a significant prevalence of coronary artery disease but no prior history of ventricular arrhythmias or sudden cardiac arrest. Participants were enrolled from several sources, including individuals transported by EMS for symptoms of acute coronary ischemia, patients undergoing angiography or attending outpatient cardiology clinics at a participating hospital, and members of a regional health maintenance organization.

### Study design

For this study, we performed a case-control study in which all cases and controls from February 1, 2002, to December 31, 2020 were selected if they were aged 40 to 75 with detailed medical records available and serum creatinine levels measured (prior to arrest for cases). If subjects had multiple serum creatinine measures, the closest to the cardiac arrest event was selected for SCA cases. For controls, the most recently measured creatinine prior to study entry was selected.

The institutional review boards of Cedars-Sinai Medical Center, Oregon Health & Science University, and all relevant hospitals/health systems have approved the study protocol.

CKD was classified based on eGFR calculated using the CKD-EPI 2021 formula and the following categories: stage 1 CKD (chart history of kidney disease, with eGFR ≥ 90 mL/min/1.73m^2^), stage 2 CKD (eGFR < 90 and ≥ 60 mL/min/1.73m^2^), stage 3a CKD (eGFR < 60 and ≥ 45 mL/min/1.73m^2^), stage 3b CKD (eGFR < 45 and ≥ 30 mL/min/1.73m^2^), stage 4 CKD (eGFR < 30 and ≥ 15 mL/min/1.73m^2^), and stage 5 CKD or end-stage renal disease (ESRD) (eGFR < 15 mL/min/1.73m^2^).^4^ Moderate CKD was defined as including both stage 3a CKD and stage 3b CKD.^4^ Demographic information and comorbidities were obtained from medical records from collaborating hospitals.

### External validation cohort selection

To validate our findings, we replicated the analysis using data from February 1, 2015, to February 1, 2023, from the ongoing Ventura PRESTO (Prediction of Sudden Death in Multi-Ethnic Communities) study, which covers a catchment population of ∼850,000. SCA cases in Ventura PRESTO were identified using methods similar to those employed in ORSUDS, including case ascertainment, adjudication, inclusion criteria, data retrieval, and definitions. The control group for Ventura PRESTO were patients from a regional health system (Cedars-Sinai Medical Center Medical Network, Los Angeles, CA) (n=4251) designed to represent members of the general population who obtain regular medical care. For the validation cohort, both cases and controls were selected using the criteria: individuals aged 40 to 75 years with detailed medical records and documented serum creatinine levels.

### Statistical analysis

Data were presented as mean and standard deviation for continuous variables and as counts with percentages for categorical variables. Tests for differences between continuous variables was evaluated using the independent samples t-test, while categorical variables were analyzed using Pearson’s chi-square test. For continuous variables with a non-normal distribution, the Mann-Whitney U test was applied to assess association. Multivariable logistic regression analysis was conducted to calculate adjusted odds ratios (ORs) for moderate kidney dysfunction as a predictor of SCA. Covariates included in the regression model were demographic factors (age, race/ethnicity, and sex) and relevant risk factors or comorbidities associated with cardiac arrest and CKD: body mass index (BMI), diabetes, hypertension, chronic obstructive pulmonary disease (COPD), liver disease, congestive heart failure (CHF), coronary artery disease (CAD), and atrial fibrillation. We also calculated the adjusted ORs for each 1 mL/min/1.73m^2^ and 10 mL/min/1.73m^2^ decline in individuals with eGFR less or equal than 90 mL/min/1.73m^2^ in discovery, validation, and pooled cohort, where the pooled cohort included all individuals from both the discovery and validation groups. Furthermore, the interaction between CKD severity and sex, age (≥ 65 vs <65), history of hypertension, diabetes, CAD, CHF in predicting SCA was also tested using the logistic regression model.

Subgroup analysis was conducted among SCA cases to highlight the relationship between CKD severity and resuscitation outcomes using multivariable regression model with covariates including demographic factors (age, race/ethnicity, and sex), history of hypertension, diabetes, CAD, CHF, witnesses, cardiopulmonary resuscitation by bystander, and initial rhythm recorded.

Initial rhythm was divided into non-shockable rhythms (pulseless electrical activity (PEA) and asystole) and shockable rhythms pulseless ventricular tachycardia and ventricular fibrillation (VT/VF). Particularly, because eGFR may vary with time, we performed a sensitivity analysis in the discovery cohort, limiting the analysis to individuals with creatinine levels measured within 90 days of the cardiac arrest event for SCA cases and within 90 days of the last physician visit for controls.

Finally, we conducted a subgroup analysis to highlight potential differences in the mechanisms linking varying severities of CKD and SCA. The cohort was stratified by CKD severity into mild/normal (eGFR ≥60 mL/min/1.73 m^2^), moderate (eGFR 30–59 mL/min/1.73 m^2^), and severe (eGFR <30 mL/min/1.73 m^2^) categories. We then compared clinical profiles and ECG characteristics between SCA cases and controls within each subgroup. All analyses were repeated for the PRESTO cohort to validate the findings. Statistical significance was determined at a two-sided p-value threshold of <0.05. All analyses were performed using IBM SPSS Statistics, version 24.

## Results

### Study overview and baseline characteristics of individuals with SCA

From February 1, 2002, to December 31, 2020, in the ORSUDS study, 6,576 SCA cases were enrolled, of which 2,068 met inclusion criteria for this analysis. Additionally, 852 controls meeting the criteria were also included (Figure 1). The mean ages of SCA cases and controls were 61.4 and 62.7 years, respectively. SCA cases exhibited a higher prevalence of risk factors and comorbidities, except for CAD, compared to controls. These included COPD, hypertension, diabetes, CHF, and atrial fibrillation (Table 1).

**Figure 1.**
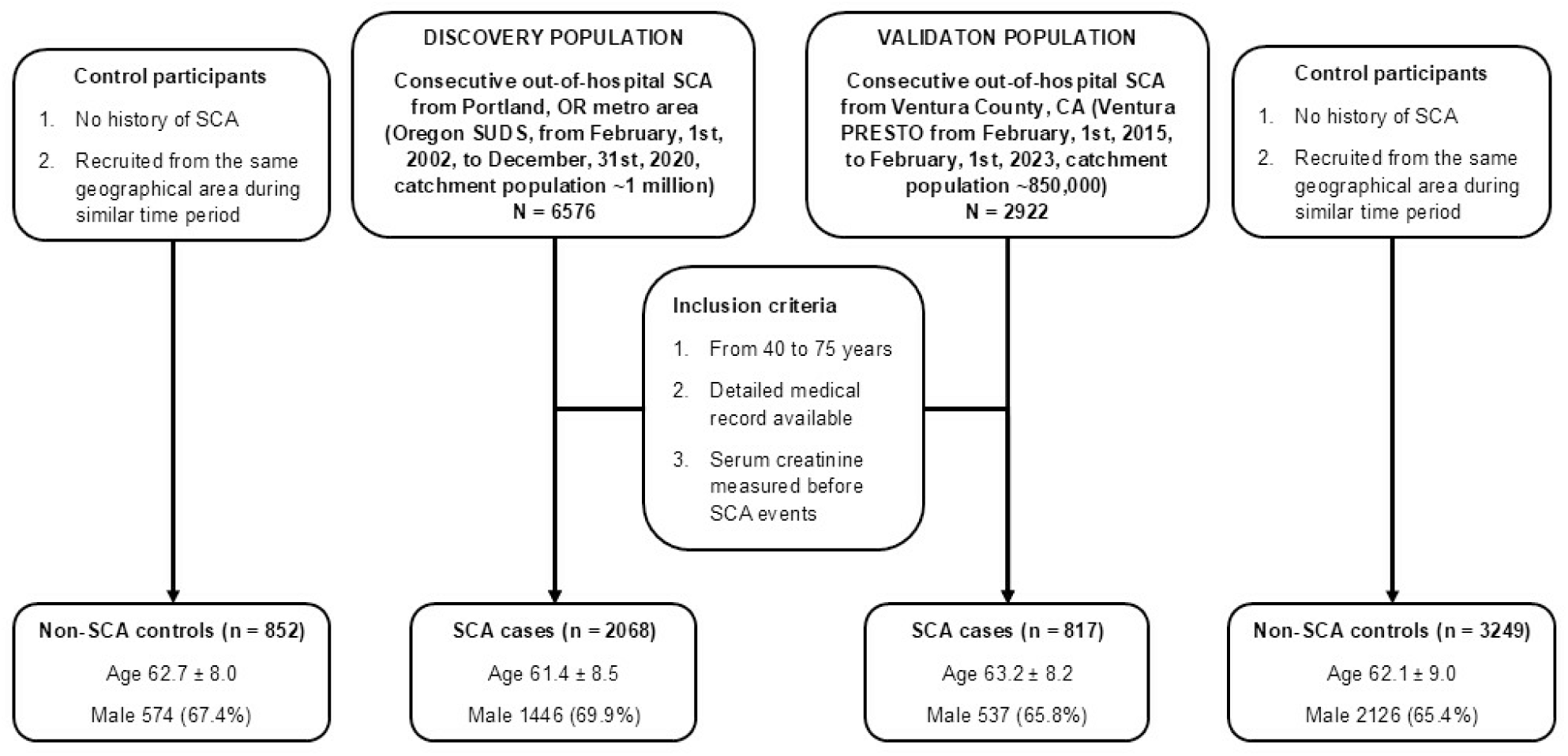
Flow chart of discovery and validation study population selection. The discovery population comprised 2,068 sudden cardiac arrest (SCA) cases and 852 non-SCA controls, aged 40 to 75 years, with detailed medical records and pre-SCA serum creatinine measurements from the Oregon Sudden Unexpected Death Study (ORSUDS). The validation population included 817 SCA cases and 3,249 non-SCA controls, aged 40 to 75 years, with detailed medical records and pre-SCA serum creatinine measurements from the Prediction of Sudden Death in Multi-Ethnic Communities (PRESTO) study in Ventura, California.

**Table 1.** Baseline characteristics of SCA cases and controls aged 40-75 in the discovery cohort (ORSUDS) from the Portland, OR metro region.

|  | SCA Cases<br>(N = 2068) | Controls<br>(N = 852) | p-value |
| --- | --- | --- | --- |
| Age, mean ± SD, year | 61.4 ± 8.5 | 62.7 ± 8.0 | <0.001 |
| Male, n (%) | 1446 (69.9) | 574 (67.4) | 0.18 |
| Race/ethnicity, n (%) |  |  | 0.02 |
| White | 1658 (81.0) | 703 (83.6) |  |
| Hispanic | 51 (2.5) | 15 (1.8) |  |
| Asian | 62 (3.0) | 17 (2.0) |  |
| African American | 225 (11.0) | 99 (11.8) |  |
| Others* | 50 (2.4) | 7 (0.8) |  |
| Missing† | 22 | 11 |  |
| eGFR, median (25th, 75th percentile), mL/min/1.73 m<sup>2</sup> ‡ | 74.7 (48.9 – 94.4) | 80.9 (66.9 – 94.1) | <0.001 |
| Chronic kidney disease by EPI2021, n (%) |  |  | <0.001 |
| Normal | 618 (29.9) | 272 (31.9) |  |
| Stage 2 | 764 (36.9) | 433 (50.8) |  |
| Stage 3 | 367 (17.7) | 125 (14.7) |  |
| Stage 4 | 129 (6.2) | 11 (1.3) |  |
| Stage 5 | 190 (9.2) | 13 (1.3) |  |
| Body mass index, mean ± SD, kg/m<sup>2</sup> | 32.7 ± 10.9 | 30.5 ± 7.0 | <0.001 |
| Chronic obstructive pulmonary disease, n (%) | 500 (24.2) | 77 (9.1) | <0.001 |
| Hypertension, n (%) | 1565 (75.8) | 600 (70.7) | 0.004 |
| Liver disease, n (%) | 258 (12.5) | 74 (8.7) | 0.004 |
| Diabetes, n (%) | 956 (46.3) | 259 (30.4) | <0.001 |
| Congestive heart failure, n (%) | 757 (36.6) | 108 (12.7) | <0.001 |
| History of coronary artery disease, n (%) | 976 (47.2) | 456 (53.5) | 0.002 |
| Atrial fibrillation, n (%) | 457 (22.1) | 114 (13.4) | <0.001 |
| SCA: Sudden cardiac arrest; SD: Standard deviation; eGFR: estimated glomerular filtration rate<br>*: Others included American Indian/Alaska Native, Native Hawaiian/Pacific Islander, and others.<br>†: For variables with missing values, proportions and p values were calculated with the non-missing data used as the denominator.<br>‡ Mann-Whitney U test |  |  |  |

### Association of moderate CKD and SCA in general population

A higher proportion of SCA cases had moderate and severe CKD (stage 4-5) compared to controls (17.7% and 15.4% vs. 14.7% and 2.6%, respectively; p < 0.001). Additionally, SCA cases exhibited a lower median eGFR than controls (74.7 vs. 80.9 mL/min/1.73m^2^, p < 0.001) (Figure 2). Using individuals with normal kidney function (stage 1) or stage 2 CKD (eGFR ≥ 60 mL/min/1.73m^2^) as the reference group, the unadjusted odds ratios for SCA were 1.49 (95% CI: 1.19–1.87) for moderate CKD and 7.39 (95% CI: 4.76–11.50) for severe CKD (stages 4–5). In models adjusted for demographic factors and comorbidities, CKD remained independently associated with risk of SCA with ORs of 1.33 (95% CI: 1.03–1.72) and 5.68 (95% CI: 3.54– 9.14) for moderate and severe CKD (Table 2), respectively. The adjusted ORs for SCA associated with other comorbidities are also presented in Table 2. Furthermore, in study participants with an eGFR ≤ 90 mL/min/1.73m^2^, each decline 10 mL/min/1.73m^2^ was associated with a 24% (OR: 1.24, 95% CI: 1.18–1.31) increase in the odds of SCA, after adjusting for demographic factors and comorbidities (Figure 3). No interaction was found between CKD severity and sex, age (≥65 years vs. <65), or history of hypertension, diabetes, CAD, or CHF in predicting the risk of SCA (Table S1). Finally, a sensitivity analysis was performed on participants with creatinine measured within 90 days of the cardiac arrest event for SCA cases and within 90 days of the last physician visit for controls. This analysis included 681 SCA cases and 278 controls. Moderate CKD remained independently associated with SCA, with a crude OR of 1.68 (95% CI: 1.18–2.38) and an adjusted OR of 1.54 (95% CI: 1.03–2.29).

**Figure 2.**
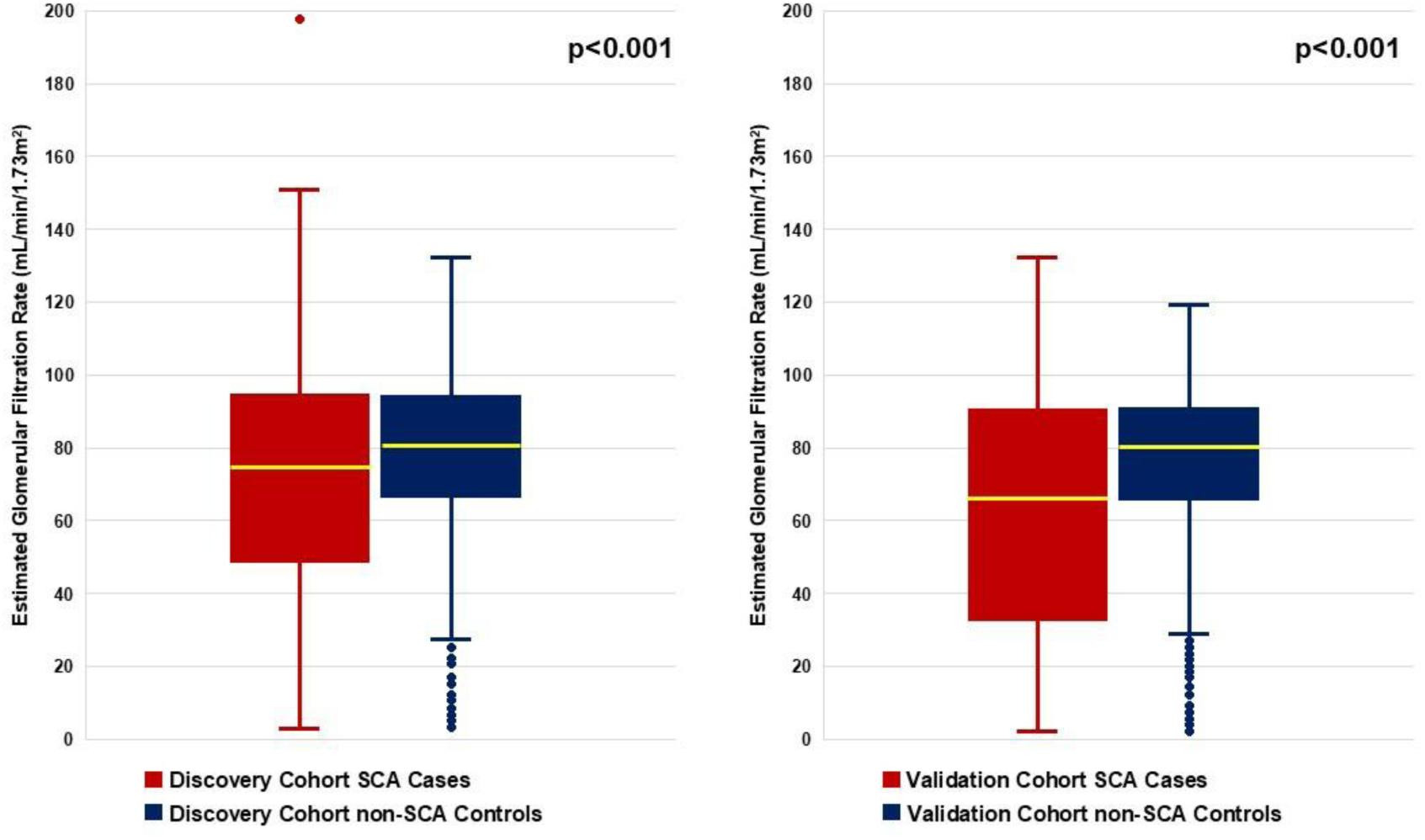
Estimated glomerular filtration rate of sudden cardiac arrest cases and controls in discovery and validation cohorts. In the discovery cohort, SCA cases had a lower median estimated glomerular filtration rate (eGFR) compared to their non-SCA control counterparts (74.7 [48.9 – 94.4] vs. 80.9 [66.9 – 94.1], p<0.001). A similar finding was observed in the validation cohort, where SCA cases exhibited significantly lower eGFR than controls (64.5 [33.0 – 90.3] vs. 78.3 [65.9 – 90.7], p<0.001).

**Figure 3.**
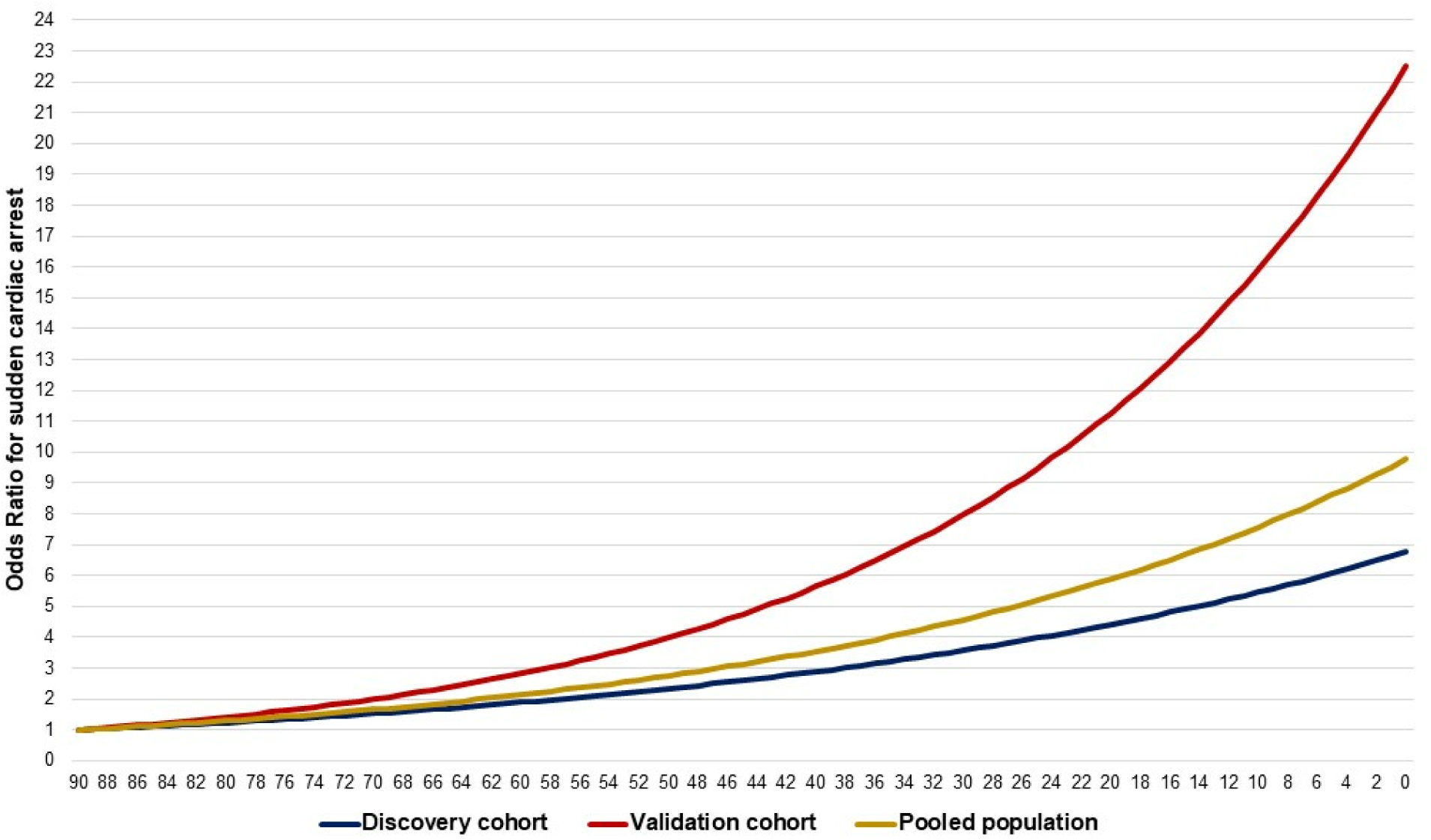
Odds ratio for risk of sudden cardiac arrest by estimated glomerular filtration rate (with ≥90 mL/min/1.73 m^2^ as reference) in discovery, validation, and pooled population. The risk of sudden cardiac arrest (SCA) increases as the estimated glomerular filtration rate (eGFR) decreases in both the discovery and validation cohorts, as well as in the pooled population.

**Table 2.** Multivariable adjusted odds ratios for sudden cardiac arrest by chronic kidney disease severity and other comorbidities in discovery (ORSUDS) and validation (PRESTO) cohorts.

|  | ORSUDS | PRESTO |
| --- | --- | --- |
| Chronic kidney disease |  |  |
| Normal or mild | Reference | Reference |
| Moderate | 1.33 (1.03 – 1.72) | 1.51 (1.17 – 1.97) |
| Severe | 5.68 (3.54 – 9.14) | 9.79 (6.33 – 15.12) |
| Chronic obstructive pulmonary disease | 3.13 (2.37 – 4.14) | 3.22 (2.35 – 4.41) |
| Hypertension | 1.04 (0.84 – 1.29) | 1.61 (1.26 – 2.05) |
| Liver disease | 1.09 (0.80 – 1.48) | 0.52 (0.38 – 0.71) |
| Diabetes | 1.41 (1.15 – 1.73) | 2.55 (2.05 – 3.18) |
| Congestive heart failure | 3.12 (2.41 – 4.04) | 3.84 (2.72 – 5.41) |
| Coronary artery disease | 0.45 (0.37 – 0.54) | 1.64 (1.29 – 2.08) |
| Atrial fibrillation | 1.22 (0.93 – 1.58) | 18.28 (11.07 – 30.19) |

### Subgroup analysis

Among SCA cases with initial rhythm data available (n=2026), individuals with normal or mild CKD were less likely to present with an initial rhythm of PEA/Asystole compared to those with moderate and severe CKD (57.4% vs. 68.7% and 69.2%, p < 0.001). Moreover, they also demonstrated a trend towards higher survival-to-hospital-discharge proportion (14.8% vs. 11.9% and 10.4%, p = 0.06) (Table 3). After adjusting for demographic factors (age, race/ethnicity, and sex), history of CAD and CHF, whether the event was witnessed, bystander-provided cardiopulmonary resuscitation, and the initial recorded rhythm, moderate CKD appeared as an independent predictor of lower survival to hospital discharge, with an adjusted OR of 0.60 (95% CI: 0.39–0.92, p = 0.02) (Table S2).

**Table 3.** Resuscitation characteristics of sudden cardiac arrest cases in discovery (ORSUDS) and validation (PRESTO) cohorts with different severity of chronic kidney disease.

|  | Discovery cohort – ORSUDS |  |  |  | Validation cohort - PRESTO |  |  |  |
| --- | --- | --- | --- | --- | --- | --- | --- | --- |
|  | Normal/mild<br>CKD<br>(n = 1382) | Moderate<br>CKD<br>(n = 367) | Severe<br>CKD<br>(n = 319) | p-value | Normal/mild<br>CKD<br>(n=446) | Moderate<br>CKD<br>(n=185) | Severe<br>CKD<br>(n=186) | p-value |
| Location, n (%) |  |  |  | <0.001 |  |  |  | <0.001 |
| Home | 866 (63.1) | 237 (64.6) | 170 (53.5) |  | 358 (80.3) | 148 (80.0) | 128 (68.8) |  |
| Care facilities† | 165 (12.0) | 66 (18.0) | 115 (36.2) |  | 38 (8.5) | 24 (13.0) | 54 (29.0) |  |
| Public | 301 (21.9) | 54 (14.7) | 28 (8.8) |  | 46 (10.3) | 12 (6.5) | 4 (2.2) |  |
| Other | 40 (2.9) | 10 (2.7) | 5 (1.6) |  | 4 (0.9) | 1 (0.5) | 0 (0.0) |  |
| Missing* | 10 | 0 | 1 |  |  |  |  |  |
| Witnessed, n (%) | 792 (57.8) | 215 (58.9) | 180 (56.6) | 0.8 | 191 (42.9) | 80 (43.2) | 71 (38.2) | 0.5 |
| Missing* | 12 | 2 | 1 |  |  |  |  |  |
| Bystander CPR, n (%) | 617 (44.6) | 141 (38.4) | 148 (46.4) | 0.06 | 240 (53.8) | 93 (50.3) | 100 (53.8) | 0.7 |
| Initial presenting rhythm, n (%) |  |  |  | <0.001 |  |  |  | 0.09 |
| VFVT | 577 (42.6) | 111 (30.8) | 98 (31.3) |  | 104 (23.3) | 39 (21.1) | 29 (15.6) |  |
| PEA/Asystole | 776 (57.4) | 249 (69.2) | 215 (68.7) |  | 342 (76.7) | 146 (78.9) | 157 (84.4) |  |
| Missing* | 29 | 7 | 6 |  |  |  |  |  |
| Survival to hospital discharge, n (%) | 204 (14.8) | 38 (10.4) | 38 (11.9) | 0.06 | 40 (9.0) | 5 (2.7) | 4 (2.2) | <0.001 |
| Missing* | 1 |  | 1 |  |  |  |  |  |
| * For variables with missing values, proportions and p values were calculated with the non-missing data used as the denominator.<br>† Care facilities includes nursing homes and outpatient clinics, emergency department<br>SCA: Sudden cardiac arrest; CKD: Chronic kidney disease; CPR: Cardiopulmonary resuscitation, VFVT: Ventricular fibrillation and ventricular tachycardia; PEA: Pulseless electrical activity |  |  |  |  |  |  |  |  |

Finally, while analyzing the differences in clinical profiles of SCA cases and controls across varying CKD severity groups, we found that COPD and BMI emerged as significant predictors of SCA in moderate CKD and normal/mild CKD but not severe CKD. The odds ratios for COPD were 3.62 (95% CI:2.61–5.02), 2.24 (95% CI: 1.19–4.24) vs. 0.93 (95% CI: 0.30–2.86), and for BMI, they were 1.014 (95% CI:1.001–1.026), 1.035 (95% CI:1.009–1.061) vs. 0.992 (95% CI:0.944–1.043) (Table S3) (Figure S1).

Regarding pre-arrest ECG parameters, which were available for 72.3% of individuals in the discovery cohort, SCA cases in both the normal/mild CKD and moderate CKD groups had significantly higher resting heart rates compared to their controls (80.3 ± 19.2 vs. 68.8 ± 14.1, p < 0.001, and 81.4 ± 19.2 vs. 69.8 ± 15.4, p < 0.001, respectively). No difference was observed in the severe CKD group (78.7 ± 17.4 vs. 78.9 ± 20.5, p = 0.96). Even after adjusting for demographics and comorbidities, resting heart rate remained a significant predictor of SCA in the normal/mild and moderate CKD groups (Table S3). Similar findings were confirmed in the validation cohort, PRESTO.

### Validation analysis

There 817 SCA cases and 3,249 controls from the PRESTO study who met the inclusion criteria, with mean ages of 63.2 and 62.1 years, respectively. As in ORSUDS, SCA cases in PRESTO had a higher prevalence of most comorbidities evaluated compared to controls. Additionally, a greater proportion of SCA cases had moderate CKD compared to controls (22.6% vs. 14.9%, p < 0.001). Using eGFR ≥ 60 mL/min/1.73m^2^ as the reference group, the crude ORs for SCA were 2.34 (95% CI: 1.92–2.85) for moderate CKD and 25.22 (95% CI: 17.94–35.45) for severe CKD (Table S4). After adjusting for demographic variables and comorbidities the adjusted ORs were 1.51 (95% CI: 1.17–1.97) for moderate CKD and 9.79 (95% CI: 6.33–15.12) for severe CKD (Table 2). Moreover, a decline in eGFR below 90 mL/min/1.73m^2^ by each 1 mL/min/1.73m^2^ and 10 mL/min/1.73m^2^ was associated to an increased adjusted OR for SCA of 1.04 (95% CI: 1.03– 1.04) and 1.41 (95% CI: 1.31–1.50), respectively.

Among SCA cases, individuals with normal or mild CKD were less likely to present with an initial rhythm of PEA/Asystole compared to those with moderate and severe CKD (76.7% vs. 78.9% and 84.4%, p=0.09). Additionally, SCA cases with normal/mild CKD demonstrated a higher proportion of ROSC and survival to hospital discharge (STHD) than their moderate and severe CKD counterparts (Table 3). After adjusting for demographic factors (age, race/ethnicity, and sex), history of CAD, CHF, witnessed status, bystander-performed cardiopulmonary resuscitation, and the initial rhythm recorded, moderate CKD appeared as an independent predictor of failure to survive until hospital discharge, with an adjusted OR of 3.19 (95% CI: 1.13–9.09, p=0.03) (Table S2).

## Discussion

We report that moderate CKD was independently associated with SCA in our discovery cohort (ORSUDS). This association was confirmed in a sensitivity analysis restricted to individuals with recently measured eGFR (within 3 months). Moreover, a reduced eGFR to below 90 mL/min/1.73m^2^ appeared to have a dose-response association with SCA. Among SCA cases, moderate CKD was associated with a higher likelihood of presenting with a non-shockable rhythm as the initial cardiac rhythm and lower survival rates to hospital discharge. These findings were validated in the geographically distinct PRESTO cohort.

Using two cohorts with distinct baseline characteristics— a younger population with a high prevalence of CAD and CHF in the discovery cohort (ORSUDS) and an older but relatively healthier population in the validation cohort (PRESTO)—, we demonstrated that moderate CKD is associated with an increased risk of SCA in the general population. This observation aligns with prior studies and underscores that the association remains significant when eGFR is estimated using either serum creatinine or cystatin C and across age groups, from younger to older individuals.^5–7,13–15,18^ Notably, we found that even a modest decline in eGFR below 90 mL/min/1.73m^2^ was associated with an increased risk of SCA, ranging from 24% to 41% for every 10 mL/min/1.73m^2^ decrease. These findings indicate a higher risk increase than previous reports, which estimated a risk increase of 11% to 17% in populations with CHF and CAD.^5,6^ This discrepancy likely reflects our study focus on the general population, which typically has a lower baseline risk of SCA than those with CHF or CAD. Consequently, the relatively greater observed impact of moderate CKD on SCA risk in our study is notable.

Moderate CKD can potentially contribute to SCA through multiple mechanisms, including exaggerated activity of the renin-angiotensin-aldosterone system (RAAS) and the sympathetic nervous system, as well as increased inflammation, fluid retention,^19^ and electrolyte imbalances.^20^ These factors collectively promote left ventricular hypertrophy, remodeling, fibrosis, and vasculopathy in both large (atherosclerosis) and small vessels (arteriolosclerosis), resulting in ischemic heart disease. These consequences not only lead to heart failure but also predispose to cardiac arrhythmias through re-entry pathways and conduction system disruption.^20–23^ As observed in our study, COPD and BMI appeared as significant predictors of SCA in individuals with normal/mild CKD and, more notably, in those with moderate CKD. This finding potentially reinforces evidence supporting the role of pathophysiological mechanisms such as inflammation and chronic hypoxia, likely intensified by the coexistence of COPD and high BMI.^24,25^ Furthermore, our study demonstrated the existence of possible enhanced sympathetic nervous system activity, as evidenced by higher resting heart rates in SCA cases compared to controls within the moderate CKD group.^26^ Therefore, prevention strategies for SCA, including pharmacological interventions such as beta-blockers, RAAS inhibitors, blood pressure and anemia management, and calcium channel blockers, should be carefully considered and initiated even in patients with a modest decline in renal function.^19,27,28^

Additionally, regarding resuscitation characteristics, among SCA cases, moderate CKD was associated with a higher proportion of initial non-shockable rhythms. These findings are consistent with our previous, machine learning based analysis of VF vs. PEA.^29^ Also these results align with previous studies on rhythm monitoring in CKD patients, particularly those undergoing hemodialysis, which showed that bradycardia and asystole, rather than ventricular arrhythmias, were the most common arrhythmias in these individuals.^30–32^ CKD patients are prone to ventricular fibrosis, which alters intracardiac conduction and is associated with conduction blocks. Furthermore, CKD patients often have anemia, which limits cell energy and makes the cardiac myocytes more vulnerable to global myocardial energy depletion, contributing to non-shockable rhythms.^29,33^ Moreover, SCA cases with moderate CKD had worse outcomes, with lower survival rates to hospital discharge compared to those with normal or mild CKD. This finding can be explained by the higher proportion of non-shockable rhythms, which were associated with poorer outcomes, and by the additional burden of CKD, which involves electrolyte and fluid imbalances.

### Strengths and Limitations

Our study possesses several strengths worth noting. Firstly, it is a community-based study encompassing two distinct populations: the ORSUDS cohort in Oregon, characterized by younger participants, a higher proportion of Caucasians, and a more prominent cardiac disease burden, and the PRESTO cohort in Ventura County, which included older participants, a higher representation of Hispanic individuals, and lower cardiac disease prevalence. Despite these demographic and clinical differences, most of our findings were consistent across cohorts.

Additionally, compared to other community-based studies, our work had a broader age range and racial/ethnic diversity, enhancing the generalizability of our findings.^7,14^ Secondly, our study design integrated data from the EMS system and a strict and systematic adjudication of SCA, providing a more comprehensive assessment and robust evaluation of the burden of SCA among CKD patients in the community compared to prior studies relying solely on ICD codes for SCA definition. Finally, our analyses incorporated adjustments for various confounders previously demonstrated to be associated with CKD and SCA, strengthening the evidence for an independent association between CKD and SCA.

However, certain limitations should be acknowledged when interpreting our findings. First, the research is based on two large community-based observational studies. While we utilized multivariable regression models to reduce confounding bias, some residual confounding may persist. Secondly, the lack of alternative methods to estimate renal function, such as eGFR calculated using cystatin C or the urine albumin-to-creatinine ratio, limits our ability to explore other perspectives of renal function and albuminuria in relation to SCA. Finally, given the nature of out-of-hospital SCA, the initial rhythm data could be influenced by delays in EMS response, potentially underestimating the proportion of shockable rhythms and limiting our ability to characterize the true underlying rhythm at the time of arrest.

## Conclusion

In conclusion, both moderate and severe CKD were associated with a significantly increased risk of SCA in the general population. Our findings underscore the importance of further research to elucidate the role of renal dysfunction in mechanisms of lethal arrhythmias and to evaluate the potential role of moderate renal dysfunction for improving SCD risk stratification.

## Acknowledgments

The authors would like to acknowledge the significant contribution of Portland, OR and Ventura, CA area residents, Global Medical Response, Portland/Gresham fire departments, local area hospital systems (Oregon Health and Science University, Kaiser Permanente, Legacy Health and Providence Health, Ventura Country Medical Center, Los Robles Regional Medical Center, Community Memorial Hospital, Simi Valley Hospital, Dignity Health Care).

## Supplemental Material

Table S1-S4 Figure S1

## Non-standard Abbreviations and Acronyms

SCD: Sudden cardiac death
SCA: Sudden cardiac arrest
CKD: Chronic kidney disease
eGFR: Estimated glomerular filtration rate
MDRD-4: Modification of Diet in Renal Disease
CKD-EPI: Chronic Kidney Disease Epidemiology Collaboration
ORSUDS: Oregon Sudden Unexpected Death Study
PRESTO: Prediction of Sudden Death in Multi-Ethnic Communities
CAD: Coronary artery disease
CHF: Congestive heart failure
COPD: Chronic obstructive pulmonary disease
PEA: Pulseless electrical activity
VT/VF: Ventricular tachycardia/Ventricular fibrillation

